# Exploring the Link Between Nitrate Exposure and Thyroid Cancer: A Nationwide State-Level Analysis

**DOI:** 10.1101/2025.10.06.25337446

**Authors:** Christina Zanazanian, Jason Semprini

**Affiliations:** Des Moines University

**Keywords:** cancer, thyroid, prevention, environment, early-onset, epidemiology

## Abstract

**Context:** Early-onset thyroid cancer incidence has been increasing, raising concerns about contributing factors.

**Objective:** We aimed to investigate the role of nitrate contamination in drinking water as a contributor to early-onset thyroid cancer incidence.

**Design:** We designed an ecological study, analyzing population-based data from the National Program of Cancer Registries. We quantified the association between nitrate exposure and early-onset thyroid cancer with a set of Poisson generalized estimating equations (GEE). Analyses adjusted for current and 7-year lagged obesity rates and access to screening.

**Setting:** Our nationwide study includes data from all 50 states in the U.S.

**Patients:** Patients between ages 0-14, 15-39, 40-59 who were diagnosed with thyroid cancer between 2003 and 2022.

**Exposures:** States were categorized into two groups based on predicted groundwater nitrate levels. States with nitrate < 2.0 mg/L were classified as “Low”, and states with levels >= 2.0 mg/L as “High”.

**Main Outcome Measures:** Population adjusted cases of thyroid cancer incidence, with patients grouped by age and sex.

**Results:** For the 0-14 or 40-59 age groups, we found no differences in thyroid incidence by nitrate exposure. For ages 15-39, there were 41.6 (CI: 6.2,77.1) more cases in high nitrate states, reflecting an 18.5% difference. Stratified by sex, in the 15-39 age group of females had 32.7 (CI: 6.3,59.1) more cases and males had 8.2 (CI: 0.1,16.4) more cases, reflecting a 17.8% and 20.4% difference respectively.

**Conclusions:** Elevated exposure to groundwater nitrate may be a significant preventable contributor to thyroid cancer in adolescents and young adults.

## Introduction

Thyroid cancer is one of the most common cancers diagnosed in young people under age sixty^1^. Early-onset thyroid cancer incidence has increased for decades, with exceptional growth in pediatric/AYA populations^2,3^. While family history, genetic mutations, and obesity are the most common drivers of thyroid cancer incidence, environmental exposures, such as radiation, also play a role^4–6^. Although underexplored, there is also a strong biological mechanistic basis for nitrate’s role in thyroid cancer etiology^7^.

Nitrate, a naturally occurring compound, is increasingly found in agricultural fertilizers, and often runs off of crop fields contaminating public and private drinking water^8^. Once consumed, nitrate acts as a competitive inhibitor of the sodium iodine symporter and prevents iodide uptake by the thyroid gland, impairing thyroid hormone synthesis^9,10^. This impairment leads to elevated levels of thyroid stimulating hormone, which can result in hypertrophy, thyroid disease, hyperplasia, and potentially malignant tumors^11–14^. Beyond hormonal impairment, nitrate consumption further contributes to carcinogenic opportunities through the reduction of nitrate to nitrite, which facilitates the formation of N-nitroso compounds which have shown to increase tumor risk in vivo^14^.

Despite the multiple potential mechanisms supporting the biological plausibility linking nitrate exposure to thyroid cancer, existing epidemiological evidence has been limited. Among the first epidemiological evaluations, a study in the agricultural state of Iowa revealed that longer exposure to higher levels of nitrate was associated with increased risk of thyroid cancer^7^. More recent evidence from California further supports the potential association between nitrate and thyroid cancer risk^15^. Notably, in both studies, the level of exposure was only half the “safe” regulatory threshold^16^. Whether the results from these two studies generalize beyond Iowa and California, and to younger age groups, remains unclear.

As nitrate levels continue to rise in our nation’s groundwater, public health systems must respond to the drinking water contamination and possible health effects of chronic, elevated nitrate consumption. Given the dearth of studies on thyroid cancer, especially in younger populations, and the expected toll of nitrate contamination on the burden of thyroid cancer^17^, our study aimed to conduct the first ever nationwide, population-based analysis quantifying the association between nitrate exposure and early-onset thyroid cancer.

## Methods

### Data and Measures

Our outcome was a state-level, annual measure of thyroid cancer incidence. Cancer incidence data were obtained from the National Program of Cancer Registries (NPCR) via the Surveillance, Epidemiology, and End Results (SEER)*Stat statistical software^18^. The NPCR provides population-based cancer incidence data covering the entire U.S. population. All data was aggregated at the state-level and included all 50 states (plus D.C.). We restricted case selection to individuals diagnosed with thyroid cancer between 2003 and 2022, ensuring twenty years of complete, non-missing data. To focus on early-onset cancer, we also restricted incidence to ages 0-59. Incidence rates were further stratified by sex and by age group, categorized as pediatric (0–14 years), adolescent and young adult (AYA; 15–39 years), and early-onset adult (40–59 years).

Our exposure of interest was a binary measure classifying states as above/below median groundwater nitrate levels. Accessing county-level groundwater nitrate data from the USGS^19^ (1991-2010), we first estimated a population weighted average of each state’s groundwater nitrate level. States with average nitrate levels < 2.0 mg/L were classified as “Low Nitrate”, whereas states with average nitrate levels >= 2.0 mg/L were classified as “High Nitrate” (Supplemental Exhibit 1).

Although the state-level analysis prohibited us from adjusting for individual-level familial or genetic thyroid cancer risk factors, our analysis did include data adjusting for two other factors contributing to both thyroid cancer risk and incident diagnoses. The first factor was obesity, which is associated with increased thyroid cancer risk^4^. Annual, age-specific obesity and overweight data were accessed from the Behavioral Risk Factor Surveillance System (BRFSS)^20^. Childhood obesity data came from State of Childhood Obesity^21^. The second factor contributing to differences in diagnosis, as opposed to risk, is screening; which we measure as time-invariant age-specific population state-level endocrinologist access^22^.

### Statistical Analysis

After reporting the observed incidence rates (cases per 100,000 population) by sex, age group, and nitrate level we conducted a series of statistical analyses to test if, after adjusting for population, obesity/overweight, and endocrinology access, thyroid cancer cases were higher in states with higher nitrate. Our primary model was a Generalized Estimating Equation (GEE) Poisson Regression model, with exchangeable correlation matrix. As an alternative sensitivity check specification, we constructed a Generalized Least Squares Random Effects Poisson Regression model. Both models estimated standard errors robust to heteroskedasticity and autocorrelation. Both models included a population exposure offset, essentially modelling differences in cases as an implicit rate differential. Both models adjusted for current and 7-year lagged obesity, as well as overweight rates, and the proportion of the age-specific population with access to an endocrinologist. In addition to estimating absolute modelled differences in cases, we estimated each model with a log-transformed dependent variable to quantify the percentage difference in cases between high and low nitrate states.

## Results

### Summary Statistics

Table 1 presents the incidence rates of thyroid cancer per 100,000 population, grouped by nitrate exposure (low vs. high). Individuals within the early-onset adult age group (40 – 59) had the highest overall incidence rate of thyroid cancer (21.42 cases per 100,000), followed by those in the age group of 15-39 (10.23 cases per 100,000). The 0 – 14 age group had the lowest overall incidence of thyroid cancer (0.31 cases per 100,000). For all age groups, overall and in both males and females, the observed incidence of thyroid cancer was higher in the high nitrate exposure states than the low nitrate exposure states. Supplemental Exhibits 2-4 report and visualize the observed, population adjusted thyroid cancer cases by nitrate group, age, and sex.

**Table 1.** Summary Statistics – Thyroid Incidence Rate (Cases per 100,000) Table 1 reports the thyroid cancer incidence rate (cases per 100,000 population) overall and by low (< 2 mg/L) and high (>= 2 mg/L) nitrate state groups. The incidence rate for sex stratified pediatric populations was suppressed due to case counts < 16.

|  | **Overall** | **Low Nitrate** | **High Nitrate** |
| --- | --- | --- | --- |
| Age 0-14 | 0.31 | 0.30 | 0.32 |
| Age 15-39 | 10.23 | 9.39 | 10.87 |
| Age 40-59 | 21.42 | 19.89 | 22.61 |
| Age 0-14 Females | SUPRESSED | SUPRESSED | SUPRESSED |
| Age 15-39 Females | 17.03 | 15.70 | 18.07 |
| Age 40-59 Females | 32.14 | 29.77 | 33.98 |
| Age 0-14 Males | SUPRESSED | SUPRESSED | SUPRESSED |
| Age 15-39 Males | 3.60 | 3.23 | 3.88 |
| Age 40-59 Males | 10.44 | 9.72 | 11.01 |

### Primary Results

Figures 1-2 visualize the population-adjusted, modelled differences in thyroid cancer cases by state nitrate level by age group and sex. Table 2 reports the modelled differences in thyroid cancer cases, between high and low nitrate exposure states. We did not find any differences between high and low nitrate states for pediatric age groups 0 to 14. For those between the ages of 15-39, adolescent/young adult (AYA) there were 41.6 (CI: 6.2,77.1) more cases for those in high nitrate states. This reflects an 18.5% difference. Stratified by sex, in the AYA group females had 32.7 (CI: 6.3,59.1) more cases and males had 8.2 (CI: 0.1,16.4) more cases, reflecting a 17.8% and 20.4% difference respectively.

**Table 2.** Population Average GEE Poisson Regression – Modelled Differences in Thyroid Cancer Cases. Table 2 reports the results of the Generalized Estimating Equation (GEE) Poisson Regression models estimating (Est.) the association between high nitrate levels (>= 2 mg/L) and thyroid cancer cases. All models adjusted for population. All regression models adjusted for current and seven-year lagged state-level rates of obesity and overweight BMI, and measures of endocrinology care access. Standard errors were estimated robust to heteroskedasticity and autocorrelation. 95% confidence intervals (CI) reported in brackets. Table 2 also reports the results of a post GEE model marginal analysis, estimating the relative difference on a 0-100 percentage scale. *p<0.05, ** p < 0.01, *** p < 0.001.

|  | **Est.**<br>**[95% CI]** | **% Difference**<br>**[95% CI]** |
| --- | --- | --- |
| Age 0-14 | 0.0<br>[-0.5, 0.5] | 0.5%<br>[-10.4%, 11.5%] |
| Age 15-39 | 41.6*<br>[6.2,77.1] | 18.5%<br>[3.7%, 33.9%] |
| Age 40-59 | 46.4<br>[-5.1,97.8] | 12.2%<br>[-1.1%, 25.4%] |
| Age 15-39 Females | 32.7*<br>[6.3,59.1] | 17.8%<br>[3.8%, 31.8%] |
| Age 40-59 Females | 33.2<br>[-3.3,69.7] | 11.5%<br>[-0.9%, 23.9%] |
| Age 15-39 Males | 8.2*<br>[0.1,16.4] | 20.4%<br>[0.4%, 40.4%] |
| Age 40-59 Males | 13.8<br>[-1.3,29.0] | 15.3%<br>[-1.3%, 31.8%] |

**Figure 1.**
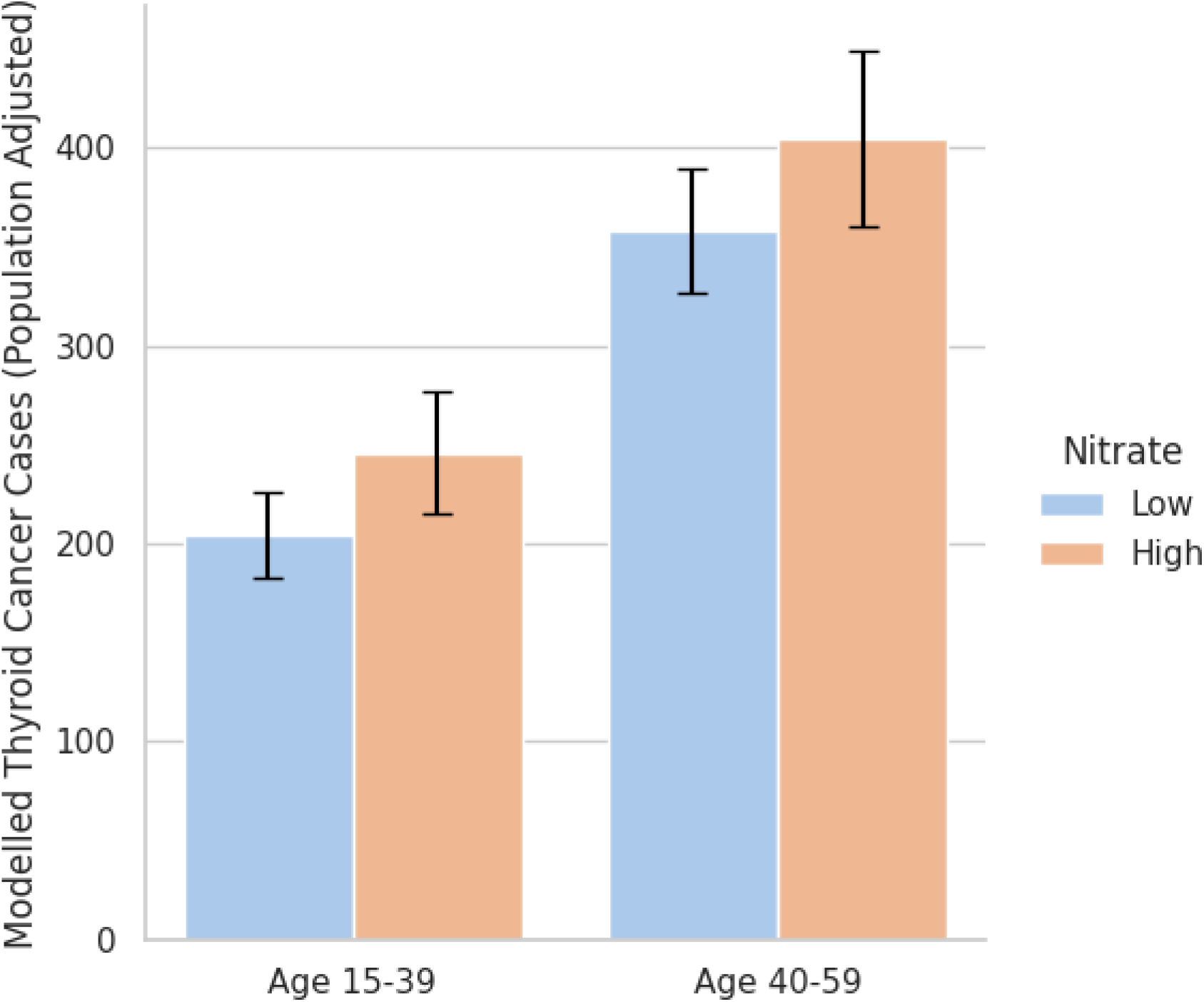
Modelled Thyroid Cancer Cases (adjusted for population) by age group and nitrate level. Figure 1 visualizes the modelled difference in population-adjusted thyroid cancer case counts from generalized estimating equation (GEE) Poisson regression models. Models adjust for state random effects and year fixed effects, as well as current and seven-year lagged state-level rates of obesity and overweight BMI, and measures of endocrinology care access. Error bars represent 95% confidence intervals. High nitrate indicates states with >= 2 mg/L average groundwater nitrate based on predicted measures.

**Figure 2.**
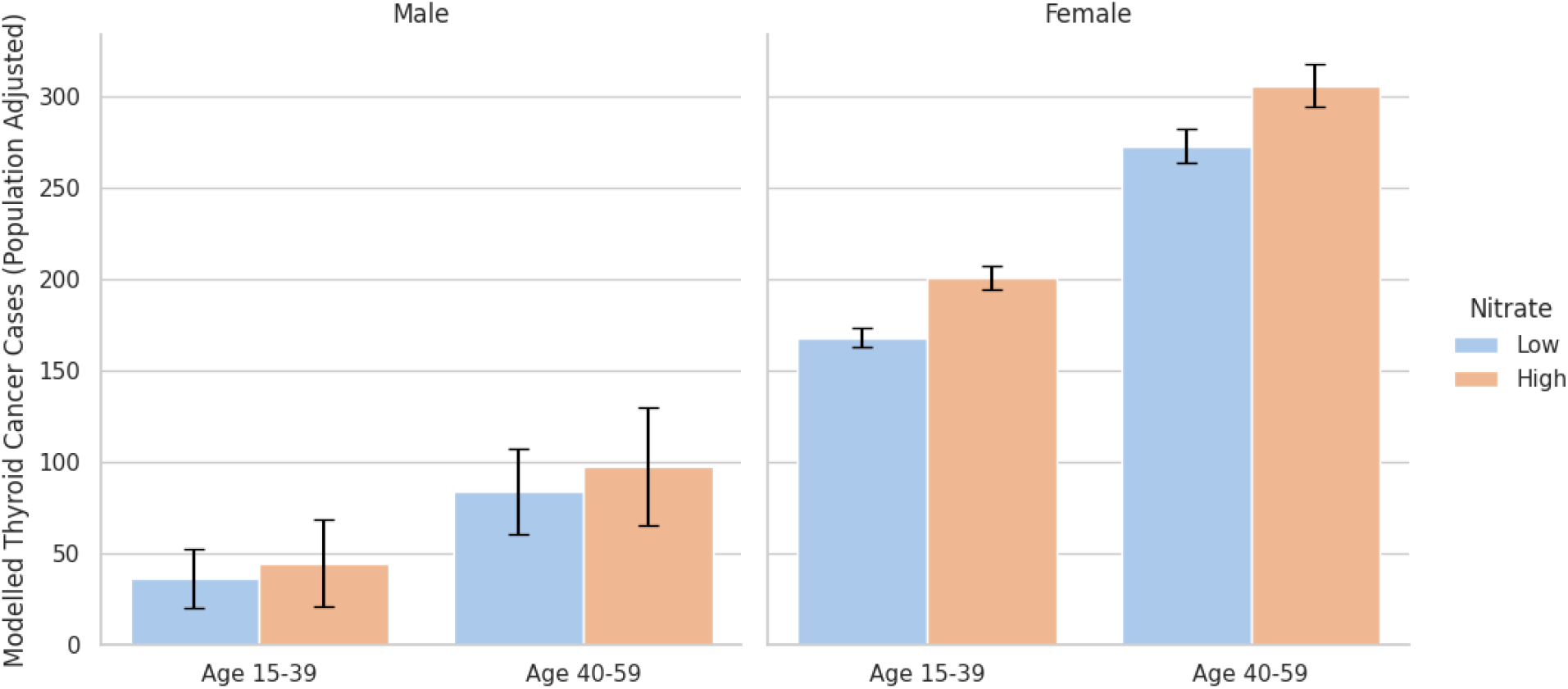
Observed Thyroid Cancer Cases (adjusted for population) by sex, age group, and nitrate level. Figure 2 visualizes the sex-stratified, modelled difference in population-adjusted thyroid cancer case counts from generalized estimating equation (GEE) Poisson regression models. Models adjust for state random effects and year fixed effects, as well as current and seven-year lagged state-level rates of obesity and overweight BMI, and measures of endocrinology care access. Error bars represent 95% confidence intervals. High nitrate indicates states with >= 2 mg/L average groundwater nitrate based on predicted measures.

Conversely, for early-onset adult populations between ages 40 and 59, there were no statistically significant differences between state nitrate groups. However, the 95% confidence interval rules out large negative differences (< −5.1 cases or <1.1%) in the high nitrate states. For AYA populations aged 15–39 years, GLS RE estimates were consistent for the overall group and in females (but not males). The GLS RE estimates (null) for the pediatric and early-onset age group were consistent with the GEE estimates.

## Discussion

With rising levels of thyroid cancer, it is important to understand how exposure to nitrate contamination in drinking water influences incidence. Our research found that thyroid cancer in adolescent and young adult populations was consistently higher in states with high groundwater nitrate exposure compared to states with low nitrate exposure. This association between nitrate exposure and thyroid cancer incidence were minimal and statistically insignificant in pediatric (< 15 years old) and adult (40-59 years old) groups. Even after adjusting for obesity and access measures, our models suggested that the largest association between nitrate exposure and thyroid cancer incidence was found in 15-39-year-old females.

These results were consistent with many preexisting studies, further suggesting a potential association between nitrate and increased thyroid cancer risk^7,15,17,23^. In addition to focusing on sex stratified, early-onset thyroid cancer incidence, our major contribution to the existing evidence base was analyzing nationwide cancer data for the entire U.S., with models adjusting for both observable heterogeneity (i.e., obesity, endocrinologists) and unobservable heterogeneity (random effects) at the state level. Although this study was not designed quantify a causal relationship; our methodology revealed convincing evidence that reducing nitrate exposure could potentially reduce early-onset thyroid cancer incidence.

To better understand any causal relationships and underlying mechanisms, future research should utilize individual-level data with quasi-experimental designs. Heterogeneity or mediation analyses could further provide the public with critical evidence for policymakers and healthcare providers by identifying which subpopulations may be at greatest risk of increased incidence of early-onset thyroid cancer with elevated exposure to nitrates in water. Additionally, studies investigating the impact of targeted interventions, such as programs or practices aimed at reducing nitrate exposure, could provide valuable insights into how such measures may influence the incidence of thyroid cancer. These future studies can help guide the development of public health strategies and policies aimed at mitigating the adverse effects of nitrates on cancer.

The emerging epidemiology evidence, backed by the well-established multifaceted biological mechanisms linking nitrates to thyroid cancer warrant, not just greater research attention to understand the role of nitrate, but greater regulatory attention to mitigate the adverse effects of nitrates in drinking water with elevated nitrate on the development of early onset thyroid cancer^7,9,10,23,24^. The current EPA drinking standard of 10 mg/L in water was set in response to methemoglobinemia, with no regards to cancer^14,25,26^. Regulatory efforts must include a portfolio of protections for public water systems as well as private well owners^27^. However, to effectively reduce the risk of early-onset thyroid cancer incidence by reducing exposure to groundwater nitrate contamination, policymakers must aim upstream by incentivizing and requiring improved agricultural nitrate reduction practices^28–30^.

## Limitations

Our study is not without its limitations. First, the analyses were observational and therefore cannot establish causality. While we identified associations between nitrate levels and thyroid cancer incidence, these results may be subject to residual confounding, reverse causation, or other sources of bias inherent to ecological studies. Second, the models used were complex and our conservative inference resulted in wide confidence intervals, potentially increasing Type II error. Third, our measure of nitrate exposure relied on a single state-level classification derived from archived county-level groundwater data (1991–2010). Although this represented the best available nationwide data, it may not accurately reflect more dynamic changes in nitrate levels or within-state heterogeneity. Because thyroid cancer incidence data were not available at the county level by age and sex, we were unable to evaluate finer geographic or subgroup variation, which could obscure more granular associations with higher nitrate exposures. Additionally, our reliance on state-level, aggregated data introduces the risk of ecological fallacy. Associations identified at the state level may not apply to individuals within those states, and therefore readers should avoid causal interpretations of our study.

## Conclusions

In summary, thyroid cancer was consistently higher in those aged 15-39 residing in high ground water nitrate states. Regulation of nitrate contamination in drinking waters at the source can be protective against the development of early onset thyroid cancer in adolescents and young adults. Future studies utilizing individual level, prospective data could help guide the development of future public health strategies and policies.

## Supporting information

Supplemental Exhibits

## Data Availability

Data Availability: All analytic data for this study is shared as a supplemental file.

https://github.com/jsemprini/thyroid_nitrate_NPCR

## Acknowledgements

Des Moines University Mentored Student Research Program

