## Supplemental Exhibits for "Exploring the Link Between Nitrate Exposure and Thyroid Cancer: A Nationwide State-Level Analysis"

**Supplemental Exhibit 1: State Groups – Nitrate Levels**

| **Low Nitrate**  **(< 2 mg/L)** | **High Nitrate**  **(> 2mg/L)** |
| --- | --- |
| Arkansas | Alabama |
| Florida | Arizona |
| Illinois | California |
| Louisiana | Colorado |
| Maine | Connecticut |
| Massachusetts | Delaware |
| Michigan | Georgia |
| Mississippi | Idaho |
| Missouri | Indiana |
| Montana | Iowa |
| New Hampshire | Kansas |
| New Mexico | Maryland |
| North Carolina | Minnesota |
| North Dakota | Nebraska |
| Oklahoma | Nevada |
| South Carolina | New Jersey |
| South Dakota | New York |
| Tennessee | Ohio |
| Texas | Oregon |
| Vermont | Pennsylvania |
| Virginia | Rhode Island |
| West Virginia | Utah |
| Wyoming | Washington |
|  | Wisconsin |

Supplemental Exhibit 1 reports each state’s grouping based on predicted groundwater nitrate levels.

**Supplemental Exhibit 2 – Thyroid Cancer Cases (Observed and Modelled)**

| **Cases** | **Nitrate** | **Age** | **Sex** | **Estimate** | **Lower** | **Upper** |
| --- | --- | --- | --- | --- | --- | --- |
| Observed | Low | Age 15–39 | Both | 193.12 | 188.94 | 197.29 |
| Observed | High | Age 15–39 | Both | 220.72 | 216.78 | 224.65 |
| Observed | Low | Age 40–59 | Both | 407.08 | 400.27 | 413.89 |
| Observed | High | Age 40–59 | Both | 459.55 | 453.16 | 465.95 |
| Observed | Low | Age 15–39 | Female | 159.82 | 156.13 | 163.51 |
| Observed | High | Age 15–39 | Female | 182.31 | 178.82 | 185.79 |
| Observed | Low | Age 40–59 | Female | 303.26 | 297.62 | 308.90 |
| Observed | High | Age 40–59 | Female | 344.08 | 338.77 | 349.40 |
| Observed | Low | Age 15–39 | Male | 33.81 | 32.13 | 35.49 |
| Observed | High | Age 15–39 | Male | 39.82 | 38.22 | 41.42 |
| Observed | Low | Age 40–59 | Male | 99.01 | 95.74 | 102.28 |
| Observed | High | Age 40–59 | Male | 111.36 | 108.30 | 114.41 |
| Modelled | Low | Age 15-39 | Both | 204.27 | 182.75 | 225.80 |
| Modelled | High | Age 15-39 | Both | 245.89 | 214.79 | 277.00 |
| Modelled | Low | Age 40-59 | Both | 358.36 | 327.11 | 389.61 |
| Modelled | High | Age 40-59 | Both | 404.72 | 359.96 | 449.48 |
| Modelled | Low | Age 15-39 | Female | 167.70 | 151.69 | 183.72 |
| Modelled | High | Age 15-39 | Female | 200.40 | 176.62 | 224.17 |
| Modelled | Low | Age 40-59 | Female | 272.50 | 249.04 | 295.96 |
| Modelled | High | Age 40-59 | Female | 305.70 | 273.36 | 338.04 |
| Modelled | Low | Age 15-39 | Male | 36.27 | 31.18 | 41.34 |
| Modelled | High | Age 15-39 | Male | 44.50 | 38.08 | 50.92 |
| Modelled | Low | Age 40-59 | Male | 83.85 | 74.58 | 93.12 |
| Modelled | High | Age 40-59 | Male | 97.68 | 85.94 | 109.42 |

Supplemental Exhibit 2 reports the observed and modelled population-adjusted thyroid cancer case counts by sex, age, and state-level nitrate grouping. Modelled cases derived from generalized estimating equation (GEE) Poisson regression models. Models adjust for state random effects and year fixed effects, as well as current and seven-year lagged state-level rates of obesity and overweight BMI, and measures of endocrinology care access.

**Supplemental Exhibit 3 – Observed, Population-Adjusted Thyroid Cancer Cases**

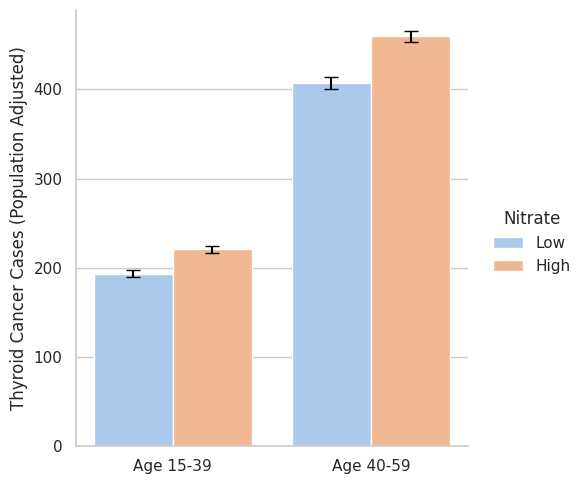

Supplemental Exhibit 3 visualizes the observed difference in population-adjusted thyroid cancer case counts from generalized estimating equation (GEE) Poisson regression models. Error bars represent 95% confidence intervals. High nitrate indicates states with >= 2 mg/L average groundwater nitrate based on predicted measures.

**Supplemental Exhibit 4 – Observed, Population-Adjusted Thyroid Cancer Cases – by sex**

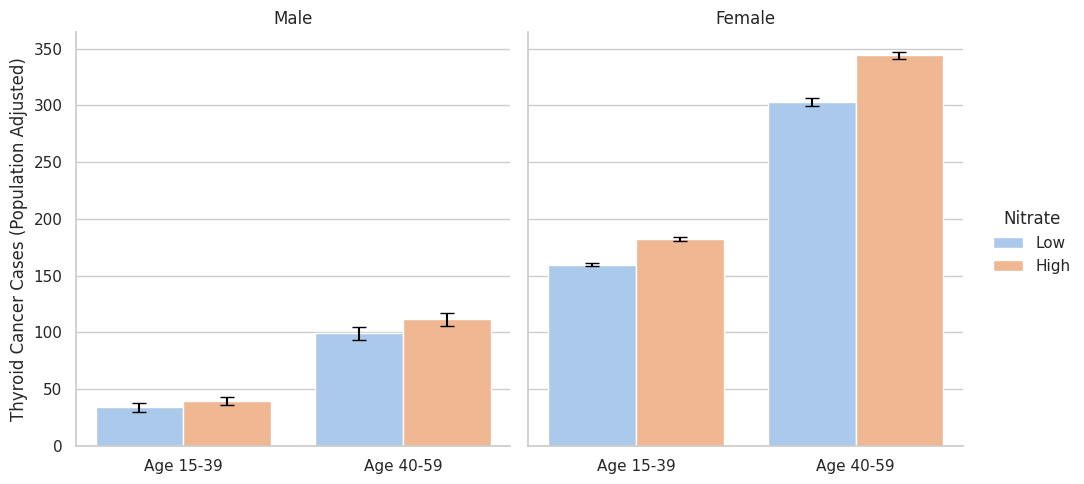

Supplemental Exhibit 4 visualizes the sex-stratified, observed difference in population-adjusted thyroid cancer case counts from generalized estimating equation (GEE) Poisson regression models. Error bars represent 95% confidence intervals. High nitrate indicates states with >= 2 mg/L average groundwater nitrate based on predicted measures.

**Supplemental Exhibit 5 –Population Average GLS RE Poisson Regression –
Modelled Differences in Thyroid Cancer Cases**

|  | **Est.**  **[95% CI]** |
| --- | --- |
| Age 0-14 | -0.2  [-0.6, 0.3] |
| Age 15-39 | 37.4*  [5.0,69.9] |
| Age 40-59 | 34.0  [-25.6,93.7] |
| Age 15-39 Females | 29.2*  [5.4,53.0] |
| Age 40-59 Females | 28.7  [-12.2,69.6] |
| Age 15-39 Males | 6.0  [-0.7,12.7] |
| Age 40-59 Males | 9.0  [-6.0,24.0] |
